# Understanding Diagnostic Uncertainty: Comparing Clinicians Pre-Test Probability of Pneumonia to Bronchoalveolar Lavage Results in Critically Ill Patients

**DOI:** 10.1101/2025.07.04.25330849

**Authors:** Caroline Zhao, Catherine Gao, Helen K. Donnelly, Erin Korth, Francisco Martinez, Bridget Giblin, Leslie Pinzon, Katie Clepp, Nandita R. Nadig, Benjamin D. Singer, Richard G. Wunderink, Chiagozie Pickens, NU SCRIPT INVESTIGATORS

## Abstract

**Rationale:** While clinical criteria are used to diagnose and treat pneumonia in critically ill patients, rates of concordance between a clinician’s suspicion for pneumonia and a confirmed diagnosis using bronchoalveolar lavage (BAL) results are undefined. Factors that contribute to diagnostic concordance, and clinical outcomes associated with diagnostic discordance, are unknown.

**Objective(s):** To assess rates of diagnostic concordance between clinicians’ pre-test probability of pneumonia and BAL-confirmed diagnosis, and to identify clinical factors and outcomes associated with diagnostic discordance in an intensive care unit (ICU) population.

**Methods:** This was a single-center, prospective observational study of intubated, mechanically ventilated patients. From 2018 to 2022, clinicians were asked to provide a pre-test probability of pneumonia on the same day they performed a bronchoalveolar lavage for the patient.

**Results:** Among 659 patients, 84% (553/659) had pneumonia. Diagnostic concordance occurred in 80% (445/553) of these cases. Clinicians assigned a low pre-test probability for pneumonia to 20% (109/553) of patients with confirmed pneumonia. Clinicians assigned a high pre-test probability for pneumonia in 28% (30/106) of patients without pneumonia. Therefore, overdiagnosis in the setting of no pneumonia occurred more often than a missed diagnosis in the setting of true pneumonia (28% vs 20%, p = 0.05). Amongst patients with pneumonia, there were no significant differences in vital signs or laboratory values between those assigned a low pre-test probability of pneumonia and those assigned a high pre-test probability of pneumonia. In patients with culture negative pneumonia (n = 117), those assigned a low pre-test probability of pneumonia, compared to those assigned a high pre-test probability of pneumonia, had a longer length of stay in the hospital (36 days vs 18 days, p = 0.02) and the ICU (21 days vs 9 days, p = 0.01).

**Conclusions:** Over-diagnosis, rather than a missed diagnosis, is the more frequent cause of diagnostic discordance. In culture-negative pneumonia, a low-pretest probability is associated with longer lengths of stay in the hospital and ICU. Future research should explore alternative approaches to improve diagnostic accuracy in critically ill patients.

## Introduction

Pneumonia is diagnosed by clinical signs and symptoms of respiratory infection with a radiographic correlate.^1^ However, these criteria can have a specificity as low as 55%, leading to high rates of discordance between the initial and final diagnoses in hospitalized patients with suspected pneumonia.^2,3,4^ Furthermore, compared to patients who have a correct initial diagnosis, patients with an incorrect initial diagnosis of pneumonia have a higher 30-day mortality.^5^ It is therefore important to identify factors contributing to diagnostic discordance. In critically ill patients who are intubated, diagnostic uncertainty in pneumonia may be heightened due to the difficulty in chest radiograph interpretation of intubated patients, variation in clinical presentation, and occurrence of other syndromes that have overlapping features.^6^ Differences between a clinician’s initial and final diagnosis may impact treatment, hospital length of stay, and decision to order additional tests. Therefore, there is a need to identify critically ill patients at risk of diagnostic discordance and understand factors that may influence a clinician’s pre-test probability of pneumonia in the intensive care unit (ICU).

Importantly, while many studies have examined diagnostic uncertainty in patients with an initial diagnosis of pneumonia, the final diagnosis of pneumonia is often ascertained from ICD-10 codes and/or clinical notes which do not accurately capture severe pneumonia or the etiology of pneumonia.^7^ Few studies of clinician diagnostic accuracy have leveraged lower respiratory tract sampling to confirm or exclude the diagnosis of pneumonia. We therefore conducted a prospective, observational study in critically ill, mechanically ventilated patients who had a BAL obtained by their clinical team. We specifically sought to define the relationship between a clinician’s pre-test probability of pneumonia and the presence of lower respiratory tract infection diagnosed by BAL and confirmed by physician adjudication.

## Methods

### Study Cohort

This study was conducted within the Successful Clinical Response to Pneumonia Therapy (SCRIPT) Systems Biology Center. SCRIPT is an NIH-funded, single-center, prospective observational study of mechanically ventilated patients with suspected pneumonia (IRB #STU00204868).^8-10^ Patients in the ICU are screened and approached for enrollment in SCRIPT if the clinical team has ordered a BAL obtained for clinical purposes. Clinical testing of all BAL samples includes semi-quantitative culture, multiplex PCR testing for common pneumonia pathogens, and cell count and differential.

### Assessment of Pre-test Probability

From July 2018 to November 2022, attending physicians or ICU fellows were approached by a member of the research team and asked to select one of the following options for a pre-BAL probability of pneumonia: <15%, 30%, 50%, 70%, >85%. This question was asked on the day the BAL was performed. The research team recorded the response of the clinician on a paper form and entered the response into an electronic data collection tool.

### Clinical Adjudication

The responses from the clinicians were compared to the adjudicated diagnosis assigned by the research team. A full description of the adjudication protocol has been published.^11^ Briefly, adjudication is performed by a panel of five ICU physicians to determine the presence and etiology of pneumonia using clinical and radiographic data, as well as results from BAL fluid testing. In the adjudication protocol, the etiology of pneumonia can be identified as ‘bacterial’, ‘viral’, ‘bacterial-viral co-infection’, ‘culture-negative’ or ‘no pneumonia.’ Note the culture-negative pneumonia is defined as a case in which no viral or bacterial organisms are detected from respiratory specimens via culture or PCR but the BAL fluid white blood cell differential contains greater than 50% neutrophils and no alternative explanation is likely.

### Outcomes of Interest and Statistical Analysis

The primary outcome of this study was the percent concordance between pretest probability and an adjudicated, BAL-confirmed diagnosis of pneumonia. We defined diagnostic concordance as the presence of pneumonia when clinicians had a high (> 50%) pre-test probability of pneumonia, or the absence of pneumonia when clinicians had a low (< 50%) pre-test probability of pneumonia. Patients with 50% pre-test probability were excluded in our analysis of concordance. We analyzed patient-level characteristics and clinical outcomes associated with diagnostic discordance. Analyses were performed using R Statistical Software (v4.3.1; R Core Team 2023). Continuous variables were reported as medians with the first and third interquartile ranges in brackets and compared using Wilcoxon rank sum test with adjustment for multiple comparisons using Bonferroni correction. Categorical variables were compared using Fisher’s Exact test and corrected for multiple comparisons. Missing data was handled via multiple imputations using the *mice* package in RStudio. The code for this project is available at https://github.com/NUSCRIPT/Pneumonia_Probability.

## Results

From July 2018 to November 2022, 659 BALs for which the pre-test probability of pneumonia was reported were available for analysis. The clinical characteristics of patients in each pre-test probability category are listed in **Table 1**. 84% (553/659) of the cases were adjudicated as pneumonia. Diagnostic concordance was present in 80% (445/553) of these cases. A low pre-test probability for pneumonia occurred in 20% (109/553) of patients with confirmed pneumonia (**Figure 1**). 16% (106/659) of cases were adjudicated as ‘no pneumonia.’ In 50% (53/106) of these cases, a low pre-test probability for pneumonia was assigned. Clinicians had a high pre-test probability for pneumonia in 28% (30/106) of the patients without pneumonia. Taken together, overdiagnosis in the setting of no pneumonia occurred more often than a missed diagnosis in the setting of true pneumonia (28% vs 20%, p = 0.05). As culture negative pneumonia can be a complex diagnosis, we separated these cases out and evaluated the concordance between pre-test probability for pneumonia in the 66% (436/659) of cases that were adjudicated as culture positive pneumonia. In 19% (82/436) of these cases, clinicians had a low pre-test probability for pneumonia, similar to the rate of discordance when culture-negative pneumonia was included. Patients with HAP were more frequently assigned a low pre-test probability for pneumonia (24.3%, 21/146) compared to patients with CAP (14.4%, 54/222) or VAP (18.4%, 34/185), (p = 0.06).

**Table 1.** Characteristics of the study cohort.

|  | **Clinician Pre-Test Probability of Pneumonia** |  |  |  |  |
| --- | --- | --- | --- | --- | --- |
|  | **15% or less**<br>N = 75<sup>1</sup> | **30%**<br>N = 87<sup>1</sup> | **50%**<br>N = 105<sup>1</sup> | **70%**<br>N = 89<sup>1</sup> | **85% or more**<br>N = 303<sup>1</sup> |
| **Pneumonia Category** |  |  |  |  |  |
| Non-pneumonia control | 30 (40%) | 23 (26%) | 23 (22%) | 14 (16%) | 16 (5.3%) |
| Community-acquired pneumonia | 10 (13%) | 11 (13%) | 18 (17%) | 20 (22%) | 87 (29%) |
| Hospital-acquired pneumonia | 21 (28%) | 33 (38%) | 36 (34%) | 32 (36%) | 100 (33%) |
| Ventilator-associated pneumonia | 14 (19%) | 20 (23%) | 28 (27%) | 23 (26%) | 100 (33%) |
| **Patient Demographics** |  |  |  |  |  |
| Age, years | 58 (48, 70) | 64 (50, 70) | 65 (56, 73) | 62 (54, 72) | 61 (50, 71) |
| Female, n (%) | 30 (40%) | 46 (53%) | 47 (45%) | 37 (42%) | 107 (35%) |
| Body Mass Index, kg/m<sup>2</sup> | 28 (25, 33) | 27 (23, 33) | 29 (24, 35) | 27 (24, 32) | 29 (25, 34) |
| Charlson Comorbidity Index | 4.0 (2.0, 7.0) | 4.0 (1.0, 7.0) | 4.0 (1.0, 7.0) | 5.0 (2.0, 8.0) | 3.0 (1.0, 6.0) |
| Immunocompromise, n (%)<sup>*</sup> | 19 (25%) | 33 (38%) | 29 (28%) | 31 (35%) | 85 (28%) |
| **Vital Signs and Labs, Day of BAL** |  |  |  |  |  |
| Temperature °F | 99.50<br>(98.80, 101.30) | 99.30<br>(98.70, 100.80) | 100.00<br>(98.78, 101.30) | 99.90<br>(98.90, 101.10) | 100.30<br>(99.10, 101.60) |
| Heart Rate, bpm | 109 (91, 127) | 108 (90, 125) | 113 (92, 128) | 104 (91, 122) | 109 (95, 128) |
| Mean Arterial Pressure, mmHg | 77 (73, 81) | 76 (72, 82) | 76 (72, 84) | 74 (71, 81) | 77 (73, 84) |
| PaO<sub>2</sub> to FiO<sub>2</sub> Ratio | 279 (173, 353) | 209 (165, 280) | 222 (166, 316) | 213 (168, 288) | 190 (148, 265) |
| Plateau Pressure, cm H<sub>2</sub>O | 21.0 (17.5, 24.7) | 23.0 (17.5, 28.3) | 21.2 (18.0, 26.8) | 21.7 (17.5, 27.5) | 24.0 (19.0, 28.7) |
| Peripheral Leukocyte Count, cells/uL | 14 (9, 21) | 13 (8, 18) | 12 (8, 17) | 13 (9, 19) | 12 (8, 19) |
| Hemoglobin, g/dL | 9.40 (8.05, 11.10) | 9.20 (7.90, 10.90) | 8.70 (7.53, 10.85) | 8.90 (8.00, 10.70) | 9.70 (8.20, 12.10) |
| Albumin, g/dL | 2.75 (2.40, 3.30) | 2.90 (2.50, 3.10) | 2.80 (2.40, 3.20) | 2.90 (2.60, 3.20) | 2.90 (2.40, 3.30) |

**Table 1.** Characteristics of the study cohort.
|  |  |  |  |  |  |
| --- | --- | --- | --- | --- | --- |
| Hospital Length of Stay, days | 22 (9, 37) | 26 (14, 46) | 26 (19, 37) | 22 (13, 40) | 24 (16, 39) |
| Total ICU Length of Stay, days | 12 (5, 27) | 19 (7, 34) | 18 (10, 29) | 13 (8, 23) | 16 (8, 30) |
| In-Hospital Mortality | 33 (44%) | 47 (54%) | 45 (43%) | 33 (37%) | 119 (39%) |

<sup>1</sup>n (%); Median (Q1, Q3)
\*Immunocompromise was defined as any condition or medication causing a patient to have a weakened immune system, including organ transplant on immunosuppressive medications, autoimmune disease, prolonged steroid usage, and malignancy.

**Figure 1.**
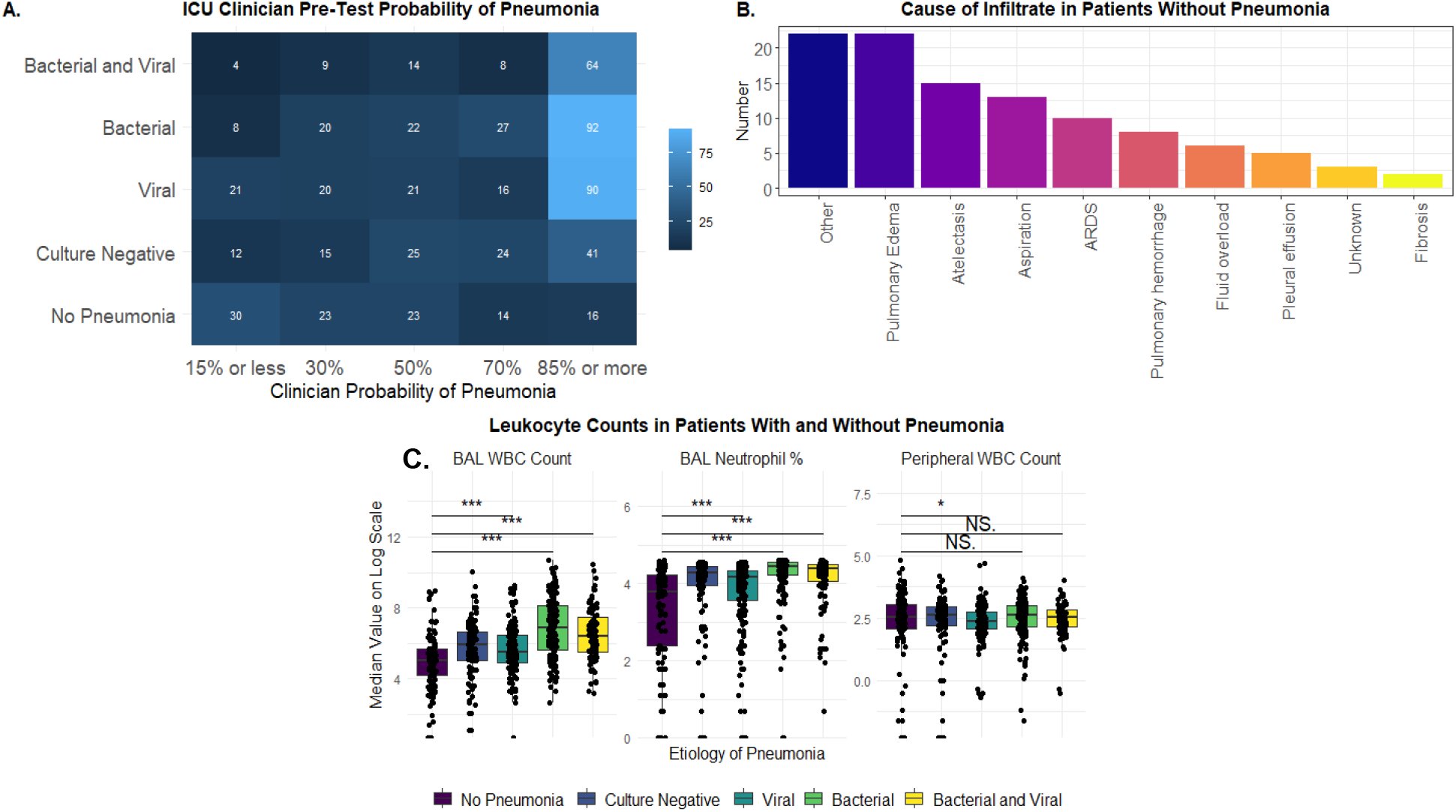
**A**. Heatmap of the pre-test probability of pneumonia and the adjudicated diagnosis. **B**. Alternative causes of infiltrates in patients without pneumonia. **C**. Leukocyte characteristics based on pneumonia etiology.

We then examined the relationship between clinical characteristics for patients who were ruled out for pneumonia (n = 106) and pre-test probability. A high pre-test probability was assigned to 30 patients and a low pre-test probability was assigned to 53 patients (23 of these patients were assigned a pre-test probability of 50%). After adjusting for multiple comparisons, there were no differences in demographics, vital signs or laboratory values between the two groups. adjusting for multiple comparisons (**Figure 2**). We also evaluated the characteristics of patients with bacterial or bacterial-viral pneumonia. A high pre-test probability was assigned to 187 patients and a low pre-test probability was assigned to 41 patients. Again, no clinical differences between patients assigned high or low pre-test probabilities of pneumonia were present. In patients with culture negative pneumonia, those who were assigned a low pre-test probability had a longer length of stay in the ICU and in the hospital compared to those assigned a high pre-test probability. Similarly, patients without pneumonia who were assigned a high pre-test probability of pneumonia had a longer length of hospital stay compared to those assigned a low pre-test probability of pneumonia (**Table 2**).

**Table 2.** Clinical outcomes of patients grouped by the pre-test probability of pneumonia and the BAL-confirmed diagnosis of pneumonia.

| Characteristic | Pre-Test Probability of Pneumonia |  |  | p-value<sup>2</sup> |
| --- | --- | --- | --- | --- |
|  | High<br>N = 30<sup>1</sup> | Intermediate<br>N = 23<sup>1</sup> | Low<br>N = 53<sup>1</sup> |  |
| Patients Without Pneumonia |  |  |  |  |
| Hospital Length of Stay, days | 23 (14, 40) | 24 (14, 32) | 19 (8, 33) | 0.2 |
| Total ICU Length of Stay, days | 14 (8, 22) | 14 (7, 25) | 7 (5, 18) | 0.026 |
| Hospital Discharge Status, n (%) |  |  |  | 0.2 |
| Alive | 19 (63%) | 9 (39%) | 32 (60%) |  |
| Died | 11 (37%) | 14 (61%) | 21 (40%) |  |
| Bacterial Pneumonia |  |  |  |  |
| Hospital Length of Stay, days | 24 (15, 42) | 29 (20, 38) | 22 (12, 44) | 0.5 |
| Total ICU Length of Stay, days | 15 (8, 32) | 18 (10, 33) | 12 (7, 28) | 0.3 |
| Hospital Discharge Status, n (%) |  |  |  | 0.3 |
| Alive | 117 (61%) | 25 (69%) | 21 (51%) |  |
| Died | 74 (39%) | 11 (31%) | 20 (49%) |  |
| Viral Pneumonia |  |  |  |  |
| Hospital Length of Stay, days | 27 (18, 38) | 31 (21, 52) | 27 (22, 47) | 0.3 |
| Total ICU Length of Stay, days | 19 (12, 31) | 21 (16, 45) | 24 (12, 40) | 0.14 |
| Hospital Discharge Status, n (%) |  |  |  | 0.12 |
| Alive | 63 (59%) | 10 (48%) | 17 (41%) |  |
| Died | 43 (41%) | 11 (52%) | 24 (59%) |  |
| Microbiology Negative Pneumonia |  |  |  |  |
| Hospital Length of Stay, days | 18 (10, 32) | 23 (14, 35) | 36 (19, 46) | 0.018 |
| Total ICU Length of Stay, days | 9 (6, 20) | 13 (8, 20) | 21 (10, 28) | 0.010 |
| Hospital Discharge Status, n (%) |  |  |  | 0.2 |
| Alive | 41 (63%) | 16 (64%) | 12 (44%) |  |
| Died | 24 (37%) | 9 (36%) | 15 (56%) |  |
| <sup>1</sup> Median (Q1, Q3); n (%) |  |  |  |  |
| <sup>2</sup> Kruskal-Wallis rank sum test; Pearson's Chi-squared test |  |  |  |  |

**Figure 2.**
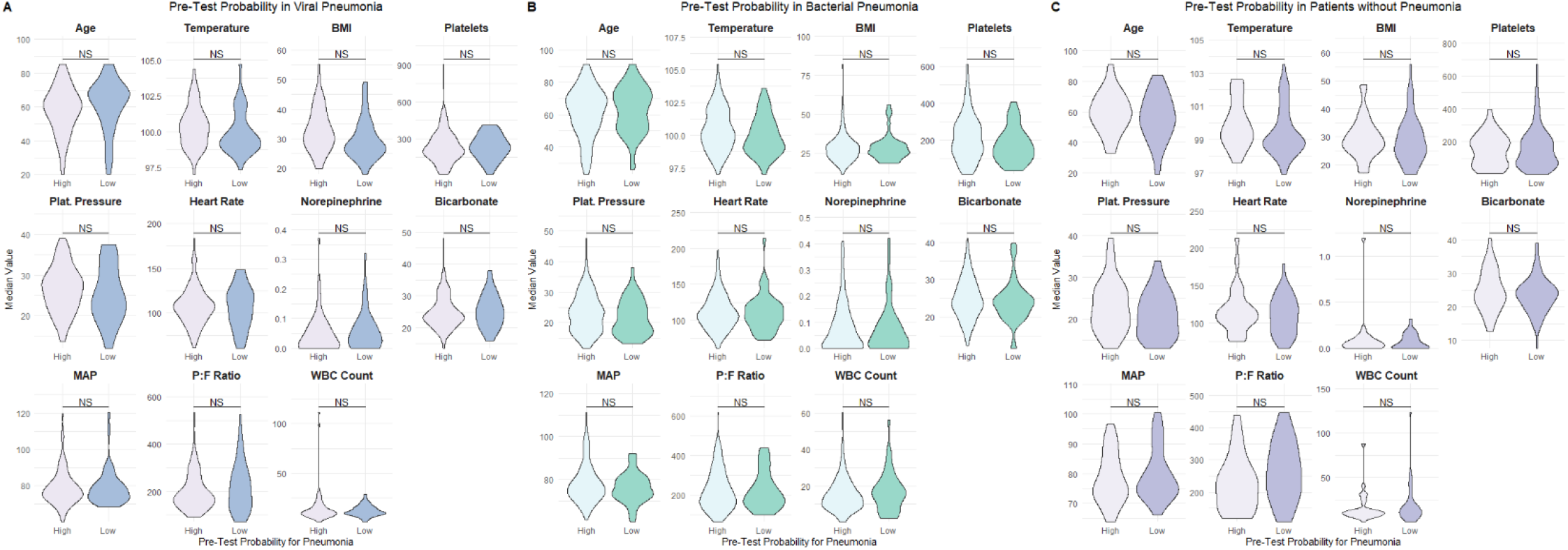
Clinical characteristics of patients who did and did not have pneumonia, grouped by clinician pre-test probability for pneumonia. When adjusted for multiple comparisons, there was no statistically significant difference in the variables between patients who were given high and low pre-test probabilities of pneumonia.

## Discussion

Our results demonstrate that in critically ill, mechanically ventilated patients, substantial discordance between the pre-test probability and final diagnosis of pneumonia exists. For the entire population, over-diagnosis of pneumonia in patients without pneumonia, compared to a missed diagnosis in patients with pneumonia, is the more common cause of discordance. This may be due to the non-specificity of clinical signs and symptoms in pneumonia in critically ill patients. Given this finding, and the associated implications for therapy (potential antibiotic overuse), additional testing with an accurate diagnostic tool like BAL should be considered in ICU patients with suspected pneumonia.

While over-diagnosis was common, surprisingly 60% of patients with a pre-test probability of 15% or less had adjudicated pneumonia. This highlights the difficulty of using clinical parameters to diagnose pneumonia, and also suggests pre-test probability for pneumonia may be influenced by variables other than routine clinical parameters. Our study draws attention to the lack of objective diagnostic tools for pneumonia. Even a single clinical sign or an abnormal radiograph has a low, but finite, association with VAP.^12^ Clinicians may overlook a diagnosis of pneumonia in patients with ventilated HAP or VAP, or those transferred to the ICU with sepsis, due to the presence of alternative sources of infection in hospitalized patients.^13^

In our cohort, 85% of patients undergoing a clinically indicated BAL and enrolled in the SCRIPT study were adjudicated to have pneumonia. This represents a higher prevalence of pneumonia compared to other studies of diagnostic accuracy in pneumonia. One potential reason for this is the routine availability of multiplex PCR for detection of respiratory viruses. In addition, our study spanned the SARS-CoV-2 pandemic. While the questionnaire did not specifically address whether the suspicion was for bacterial vs viral or other causes of pneumonia, most clinical indications for BAL in patients with a known viral respiratory tract infection was to rule out bacterial superinfection.

We add to the existing literature by demonstrating that a low suspicion for pneumonia in patients adjudicated to have culture negative pneumonia is associated with longer lengths of hospital and ICU stays. Culture negative pneumonia is a complex diagnosis, and the optimal treatment is unknown. It is possible that despite our adjudication, some of these patients had non-infectious etiologies of respiratory failure that were not responsive to antibiotics nor easily reversible, leading to a longer ICU and hospital stay. In contrast, in patients without pneumonia, a low pre-test probability of pneumonia compared to patients assigned an intermediate or high pre-test probability of pneumonia, was associated with a shorter ICU length of stay. This patient group may represent those with a clear, alternative diagnosis which could be associated with earlier initiation of appropriate management.

This study has limitations. First, the time-period of the study included the SARS-CoV-2 pneumonia pandemic which likely impacted pre-test probabilities of pneumonia. It is possible that without the presence of a pneumonia pandemic, clinical suspicion for pneumonia would be lower in some patients. However, this does not explain the 20% of cases in which clinicians had a low pre-test probability for pneumonia in patients with pneumonia. A second limitation is that this was a single center study, which limits its generalizability to other patients and institutions. In addition, clinical estimates were made at the time of BAL performance, potentially leading to a low estimated probability since a highly diagnostic test was pending and no clinical decision based on that estimate was required.

Despite these limitations, our study highlights the insufficiency of diagnosing pneumonia in critically ill patients with clinical factors alone. As such, if circumstances allow for it, our findings support BAL as a confirmatory test for the diagnosis of pneumonia in critically ill patients, to both avoiding a missed diagnosis and ensuring appropriate treatment. Future studies should evaluate adjunctive tests, biomarkers and assessments that can add accuracy to the clinical diagnosis of pneumonia in ICU patients.

## Data Availability

All data produced in the present study are available upon reasonable request to the authors.

